# Designing a Library of Lived Experience for Mental Health: integrated realist synthesis and experience-based co-design study

**DOI:** 10.1101/2023.09.06.23295134

**Authors:** Paul Marshall, Fiona Lobban, John Barbrook, Grace Collins, Sheena Foster, Zoe Glossop, Clare Inkster, Paul Jebb, Rose Johnston, Hameed Khan, Christopher Lodge, Karen Machin, Erin E. Michalak, Sarah Powell, Samantha Russell, Jo Rycroft-Malone, Mike Slade, Lesley Whittaker, Steven Jones

## Abstract

**Objective:** Living Library ‘Readers’ can learn about experiences of others through conversations with living ‘Books’. This study sought to generate a realist programme theory and a theory-informed implementation guide for a Library of Lived Experience for Mental Health (LoLEM).

**Design:** Integrated realist synthesis and experience-based co-design.

**Setting:** Ten online workshops with participants based in the North of England.

**Participants:** Thirty-one participants with a combination of personal experience of using mental health services, caring for someone with mental health difficulties, and/or working in mental health support roles.

**Results:** Database searches identified 30 published and grey literature evidence sources which were integrated with data from 10 online co-design workshops. The analysis generated a programme theory comprising five context-mechanism-outcome (CMO) configurations. For Readers, direct conversations humanise others’ experiences (CMO 2) and provide the opportunity to flexibly explore new ways of living (CMO 3). Through participation in a Living Library, Books may experience personal empowerment (CMO 4), while the process of self-authoring and co-editing their story (CMO 5) can contribute to personal development. This programme theory informed the co-design of an implementation guide highlighting the importance of tailoring event design and participant support to the contexts in which LoLEM events are held.

**Conclusions:** The LoLEM has appeal across stakeholder groups and can be applied flexibly in a range of mental health-related settings. Implementation and evaluation are required to better understand the positive and negative impacts on Books and Readers.

**Registration:** PROSPERO CRD42022312789

**Strengths and limitations of this study:**

- This study used a novel, iterative, and creative approach to integrating theory development and intervention co-design.
- The programme theory provides a conceptual basis for further evaluation of a LoLEM, including outcomes for those participating in events as ‘Books’ and ‘Readers’.
- This study informed detailed, co-designed implementation guidance for use by LoLEM organisers.
- However, the LoLEM is yet to be delivered as a sustained programme of events, so long-term impacts require further investigation.

## INTRODUCTION

The value of sharing health related experiences is widely recognised. Varied contexts draw on these experiences, including for shaping research [1], enriching professional education [2], and informing peer support [3]. There is an expanding evidence-base focused specifically on mental health lived experiences. For example, social contact interventions reduce mental health stigma [4], mental health peer support contributes to improvements in psychosocial outcomes [5], and there is growing interest in the use of mental health narratives to achieve a range of organisational aims [6]. The importance of integrating lived experience perspectives in health service development is acknowledged in UK and global policy [7,8], emphasising the need for continued efforts to promote lived experience perspectives.

Existing models for sharing mental health experiences do have limitations. Many opportunities are offered only periodically by health services and educational institutions, such as those linked to service improvement [9] or health professional training [10]. Others require commitment to more formal positions, including the peer-support worker role in the UK [11], which may preclude access by those with existing commitments. There is an outstanding need to extend such opportunities to groups whose voices are often not heard in mainstream conversations about mental health, including those from marginalised communities [12].

Adaptable approaches are required for expanding opportunities to share and learn from mental health experiences. The Living Library may represent one such model. Pioneered by the Human Library Organisation [13], it draws on a library metaphor to invite people, or ‘Readers’, to learn directly from living ‘Books’, who are supported to discuss important aspects of their life experience in short conversations [14]. Events are facilitated by staff, or ‘Librarians’, who provide guidance to those involved. These events are often held in open settings, including public and university libraries, and have typically aimed to challenge a range of societal prejudices by facilitating interactions between people who may not otherwise engage in conversation [15]. The model has also been used in a mental health context to address stigma [16] and facilitate peer-support [17]. However, implementation recommendations sensitive to the specific challenges of discussing mental health experiences are limited.

Following UK complex intervention development guidance calling for greater emphasis on theory development and stakeholder engagement [18], we applied a novel integration of realist synthesis and experience-based co-design (EBCD) to explore the Living Library as a strategy for sharing mental health experiences. Realist synthesis is a method of evidence synthesis used to develop causal statements, or programme theories, to explain how social programmes work, for whom and under which circumstances [19]. EBCD identifies opportunities for healthcare innovation and draws on stakeholder perspectives to generate creative solutions [20]. Our integrative approach is detailed in a published research protocol [21].

### Objective

- To use realist synthesis to develop a programme theory for a mental health-focused Living Library, which we term a Library of Lived Experience for Mental Health (LoLEM).
- To use insights from this synthesis to inform co-design workshops with a range of mental health stakeholders, with the goal of developing an accessible implementation guide.

## METHOD

This study received ethical approval from Coventry and Warwick National Health Service Research Ethics Committee (ref: 305975). The systematic search strategy was developed with an information specialist (JB) and pre-registered on PROSPERO https://www.crd.york.ac.uk/prospero/display_record.php?RecordID=312789. RAMESES guidance informed the reporting of this study [22].

As described in the study protocol [21], theory development and EBCD workstreams were iterative and ran in parallel. However, the study broadly progressed through the following stages:

### Eliciting initial theories and touchpoints

Initial programme theories (IPTs) were elicited through theory gleaning interviews [23] with 6 members of an expert advisory group (CI, EM, HK, MS, SF, SP) and the study public and patient involvement (PPI) lead (CL), all of whom had been involved in participating in or researching Living Libraries. Twenty IPTs were refined through advisory group feedback (appendix A). IPTs highlighted key issues to be explored with participants in co-design workshops, referred to as ‘touchpoints’ in the EBCD process [20].

### Retrieving relevant evidence

Searches were conducted on research databases and grey literature sources (figure 1) from 2000, the inception of the initial Human Library approach [13], to March 2023. Preparatory unsystematic searches indicated that relatively few articles in this literature were likely to focus solely on mental health. Therefore, reports on Living Libraries related to specific topics including mental health and from generalist events featuring a range of lived experiences were included. Following title and abstract screening, the full-texts of identified articles were judged for inclusion using a realist-informed assessment of relevance and rigour (PM) with 20% independently assessed by a second reviewer (RJ). Citation chaining was used to identify articles in reference sections and ‘cited by’ pages of articles included in the initial search.

**Figure 1.**
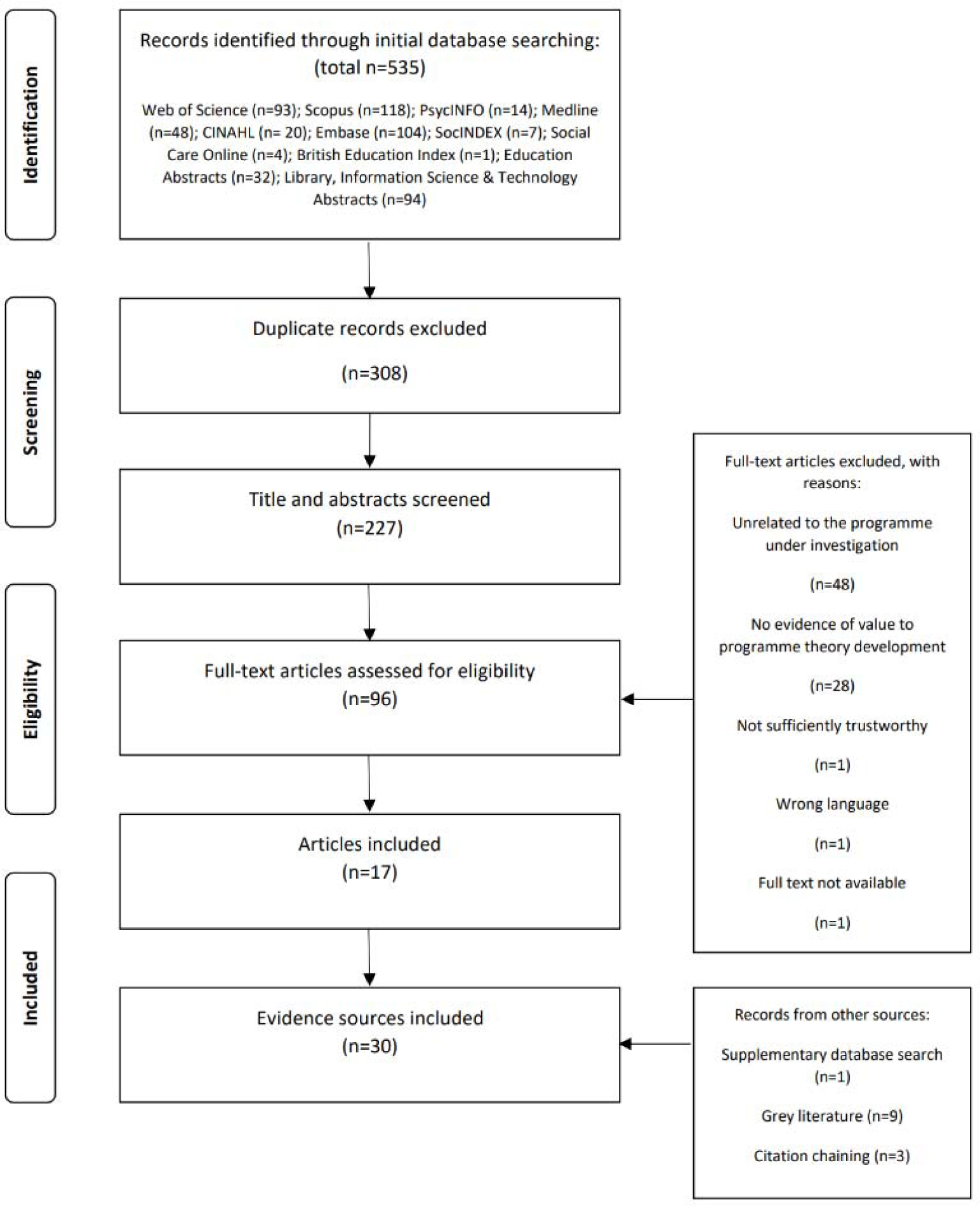
Flow diagram of evidence identification.

IPTs and initial workshops highlighted psychological safety as a key theoretical and implementation focus for this study. Consistent with the iterative nature of realist synthesis searches [24], we conducted a targeted search of a large multidisciplinary database (appendix B) for existing formal theories related to the concept and included an additional evidence source [25].

### Iterating initial theories

IPTs were added to NVivo 12 as nodes to create an initial theory framework [26]. PM conducted coding of the full text of each evidence source. This involved identifying and labelling sections of text related to potential contextual factors, mechanisms, and/or outcomes, as defined in realist methodology [19]. Codes were linked to the most relevant initial programme theory node. PM, FL, and RJ refined the initial theory framework by reformulating, consolidating, and combining initial statements based on whether underlying data supported, refuted, or refined individual IPTs. While reviewing this evidence, retroductive reasoning was used to hypothesise causal forces behind regularities apparent in the data. An iterative process of written feedback and group discussion led to the consolidation of the initial framework into a series of candidate CMO configurations.

### Establishing a multi-stakeholder co-design group

Eligible EBCD participants were adults with any self-identified mental health experience. We recruited broadly through local mental health services, health research networks, and third sector organisations primarily based in the North of England. Workshops were facilitated by KM and GC, both experts in mental health peer-support who deliver workshops from a lived experience perspective. Workshops were supported by researchers (ZG, RJ, PM), senior nurses (PJ, LW), a service user researcher (CL), and academic clinical psychologist (SJ).

### Designing workshops reflexively

We ran 10, 2-hour online co-design workshops over 12 months. Workshops were flexibly designed in team meetings around two primary goals. First, we aimed to develop a comprehensive implementation guide. This required the team to reflect on gaps in knowledge and plan further exploration of topics pertinent to implementation. Second, we explored ‘touchpoints’ to facilitate programme theory refinement. One such example was the exploration of psychological safety in workshop 4, a key concept in the IPT framework which required further investigation to understand relevant implantation factors.

### Developing and refining outputs

Workshop participants’ views and implementation suggestions were captured using the collaborative online note taking platform Jamboard [27] and in researcher (CL, PM, RJ, SJ, ZG) field notes. Data were combined in workshop summary documents following each session, reviewed by the research team in post-workshop de-briefs, and added to NVivo for integration into the ongoing analysis.

Summary documents informed an initial draft implementation guide, refined through feedback from the EBCD group. Implementation recommendations from the co-design process were linked to the developing theoretical framework. CMOs were further refined through discussion and written feedback from the wider research team and expert advisory group.

Final versions of the LoLEM programme theory and implementation guide were shared during an interactive dissemination event at which the LoLEM was piloted and the guide made freely available [28].

## RESULTS

### Results of systematic searches

Database and grey literature searches returned 30 eligible evidence sources. Figure 1 describes the process of evidence identification. Study characteristics are available in appendix C.

### Description of co-design workshops

EBCD workshops were attended by 31 participants (P). Demographic details are presented in Table 1.

**Table 1.**
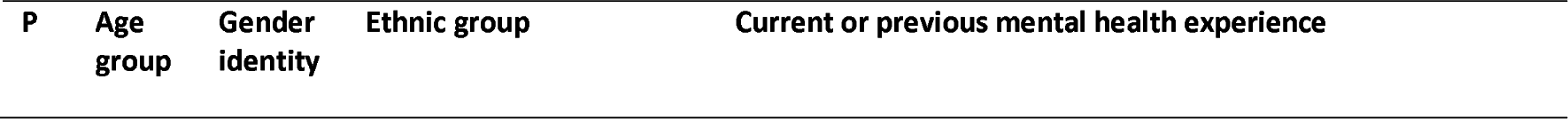
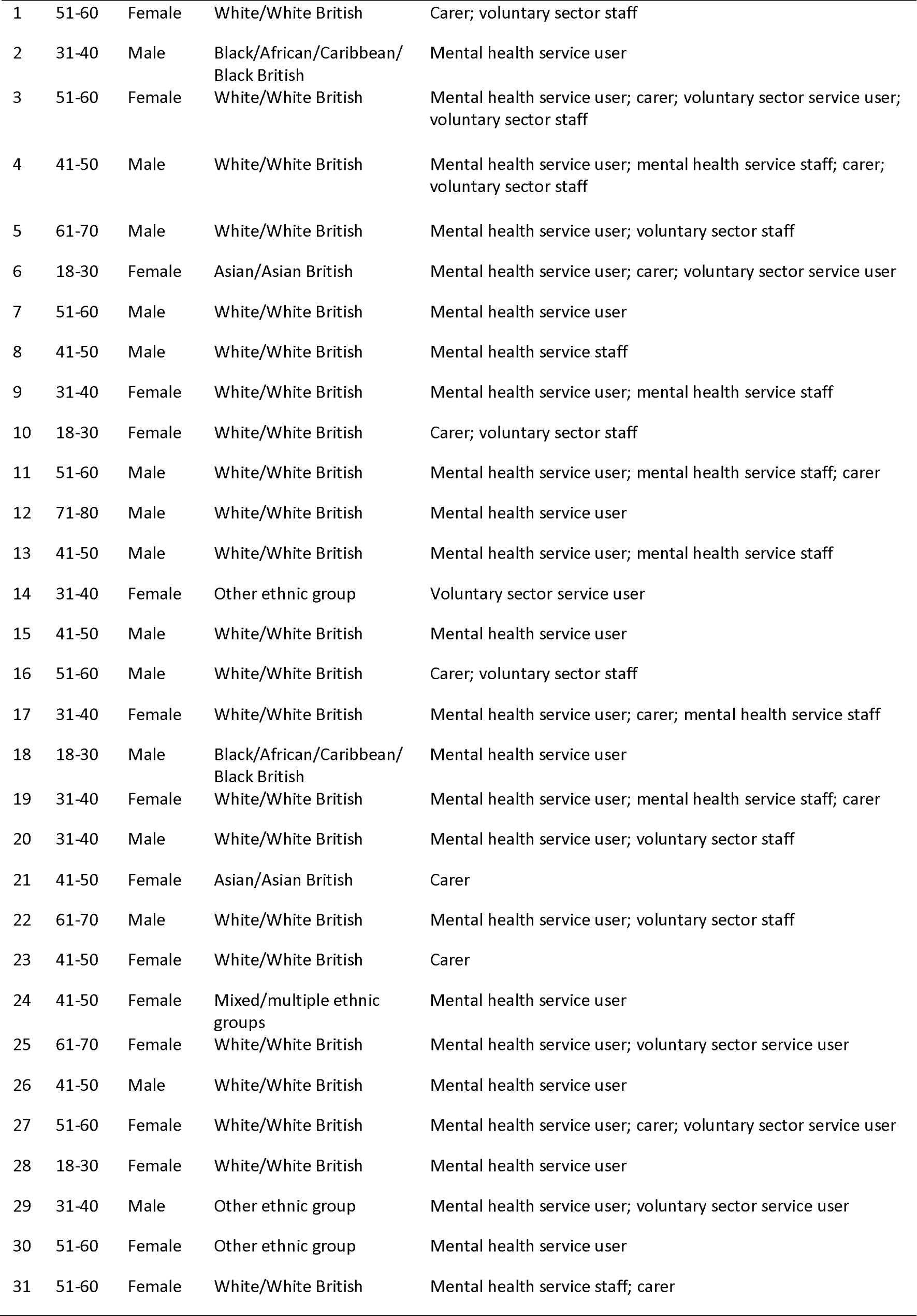
Participant demographics.

| P | Age group | Gender identity | Ethnic group | Current or previous mental health experience |
| --- | --- | --- | --- | --- |
| 1 | 51-60 | Female | White/White British | Carer; voluntary sector staff |
| 2 | 31-40 | Male | Black/African/Caribbean/<br>Black British | Mental health service user |
| 3 | 51-60 | Female | White/White British | Mental health service user; carer; voluntary sector service user;<br>voluntary sector staff |
| 4 | 41-50 | Male | White/White British | Mental health service user; mental health service staff; carer;<br>voluntary sector staff |
| 5 | 61-70 | Male | White/White British | Mental health service user; voluntary sector staff |
| 6 | 18-30 | Female | Asian/Asian British | Mental health service user; carer; voluntary sector service user |
| 7 | 51-60 | Male | White/White British | Mental health service user |
| 8 | 41-50 | Male | White/White British | Mental health service staff |
| 9 | 31-40 | Female | White/White British | Mental health service user; mental health service staff |
| 10 | 18-30 | Female | White/White British | Carer; voluntary sector staff |
| 11 | 51-60 | Male | White/White British | Mental health service user; mental health service staff; carer |
| 12 | 71-80 | Male | White/White British | Mental health service user |
| 13 | 41-50 | Male | White/White British | Mental health service user; mental health service staff |
| 14 | 31-40 | Female | Other ethnic group | Voluntary sector service user |
| 15 | 41-50 | Male | White/White British | Mental health service user |
| 16 | 51-60 | Male | White/White British | Carer; voluntary sector staff |
| 17 | 31-40 | Female | White/White British | Mental health service user; carer; mental health service staff |
| 18 | 18-30 | Male | Black/African/Caribbean/<br>Black British | Mental health service user |
| 19 | 31-40 | Female | White/White British | Mental health service user; mental health service staff; carer |
| 20 | 31-40 | Male | White/White British | Mental health service user; voluntary sector staff |
| 21 | 41-50 | Female | Asian/Asian British | Carer |
| 22 | 61-70 | Male | White/White British | Mental health service user; voluntary sector staff |
| 23 | 41-50 | Female | White/White British | Carer |
| 24 | 41-50 | Female | Mixed/multiple ethnic<br>groups | Mental health service user |
| 25 | 61-70 | Female | White/White British | Mental health service user; voluntary sector service user |
| 26 | 41-50 | Male | White/White British | Mental health service user |
| 27 | 51-60 | Female | White/White British | Mental health service user; carer; voluntary sector service user |
| 28 | 18-30 | Female | White/White British | Mental health service user |
| 29 | 31-40 | Male | Other ethnic group | Mental health service user; voluntary sector service user |
| 30 | 51-60 | Female | Other ethnic group | Mental health service user |
| 31 | 51-60 | Female | White/White British | Mental health service staff; carer |

Details of activities completed at each workshop are presented in Table 2.

**Table 2.** Experience-based co-design workshop details.

| Workshop | n | Activities |
| --- | --- | --- |
| 1 | 25 | • Project introduction<br>• Story formulation activity |
| 2 | 27 | • Story sharing activity<br>• Reflection on personal boundaries |
| 3 | 25 | • Creating a fictional Reader ‘persona’<br>• Reflection on Library design for different Reader groups |
| 4 | 24 | • What influences potential Readers’ decisions to participate?<br>• What constitutes a safe space for sharing mental health experiences?<br>• How can event organisers cultivate psychological safety amongst Books and Readers? |
| 5 | 25 | • Reflection on participants’ preferred roles at a Living Library (Books, Readers, or Librarians)<br>• Planning a hypothetical Living Library<br>• A ‘design to fail’ activity, considering factors likely to contribute to unsuccessful events |
| 6 | 23 | • Design considerations for Living Libraries in different contexts (the voluntary, the NHS, and public libraries) |
| 7 | 23 | • Identifying resources required to run successful Living Libraries<br>• A ‘sales pitch’ activity to potential funders |
| 8 | 21 | • Advantages and disadvantages of delivering Living Libraries with different aims (mental health support versus changing public perception of mental health)<br>• Co-design of the implementation guide contents page |
| 9 | 20 | • The desired characteristics of event staff (‘Librarians’)<br>• Considering how to evaluate a Living Library<br>• Feedback on the implementation guide |
| 10 | 17 | • Celebration event<br>• Review of project successes and thanking participants for their contributions |

### LoLEM programme theory

We developed a programme theory comprising five CMO configurations, with corresponding implementation recommendations (Figure 2).

**Figure 2.**
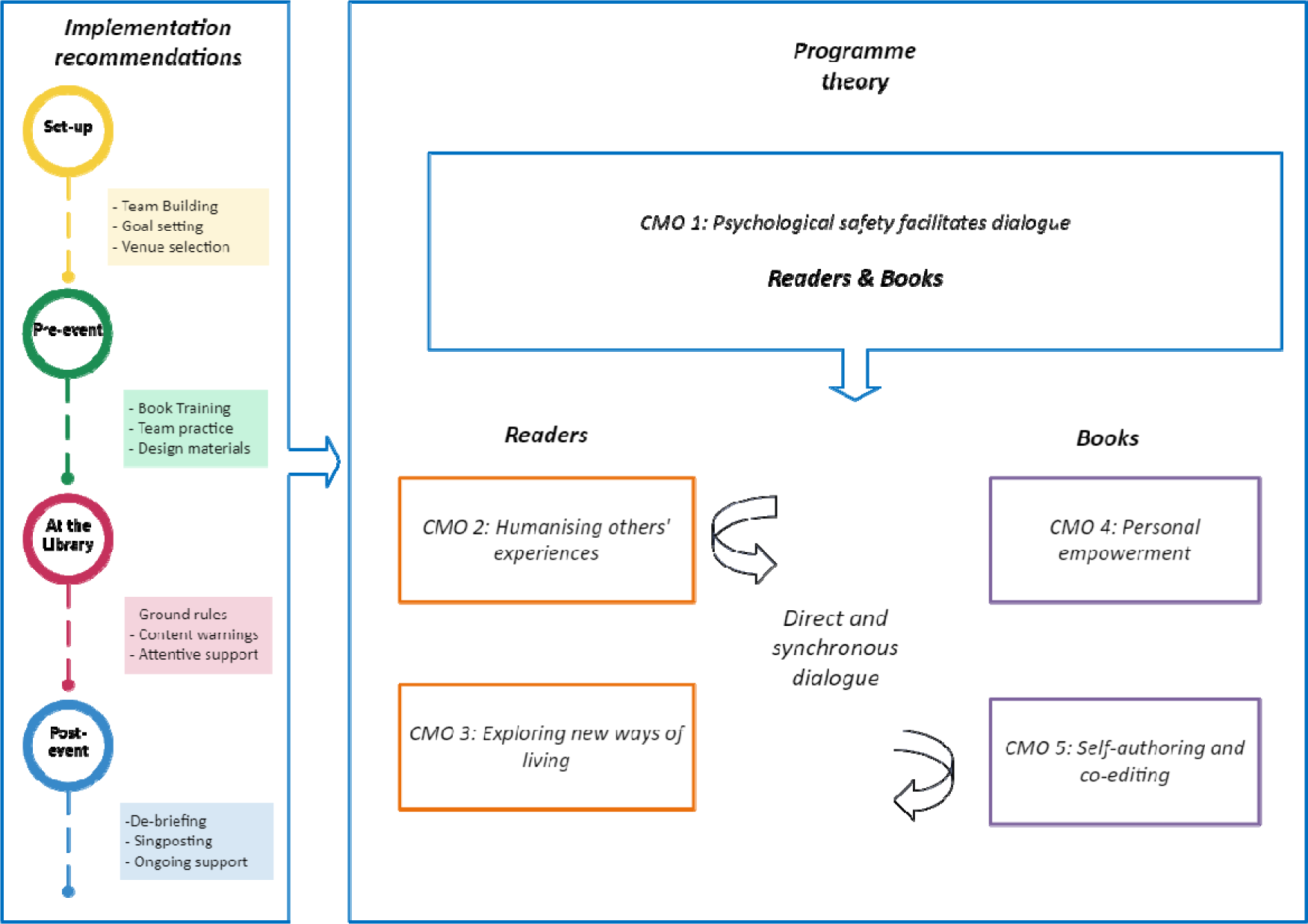
LoLEM programme theory with implementation recommendations.

### CMO 1 – Psychological safety facilitates dialogue

When event organisers implement guidance and support strategies tailored for their organisational settings (context), participants will be better able to understand the Living Library model and how to manage personal boundaries such that they feel in control of their experience (mechanism). This will promote participants’ perceptions of psychological safety (outcome) necessary for engaging and productive conversations.

Psychological safety refers to an individual’s comfort with taking interpersonal risks to meet their goals within a given organisational context [25]. The Living Library model provides a platform for participants to take interpersonal risks by sharing experiences in direct conversation. We propose that psychological safety creates a facilitating context, or ‘ripple’ effect [29], promoting the likelihood of subsequent positive impacts described by CMOs 2-5.

Developing psychological safety for Books and Readers commonly involves implementing event rules to explicitly promote mutual respect, awareness of boundaries, and a sense of personal control. Pre-event briefings may be used to reinforce the principles of the approach: “‘Readers are required to return the ‘Book’ in the same psychological and physical condition and are asked: ‘Never harm a Book!’” [30]. Promoting psychological safety for Books can involve offering training and practice sessions to set expectations, identify what they wish to share, and prepare for questions they may not want to answer. EBCD participants suggested that psychological safety would further be promoted by the reassuring presence of attentive staff, or ‘Librarians’, able to provide guidance, oversight, and emotional support where necessary.

Given their conversational focus, Living Libraries hold potential for Books and Readers to experience interpersonal challenges that could undermine psychological safety. Books may be exposed to negative attitudes towards mental health, feel pressure to be an exhaustive authority on the issues they discuss, ‘pumped’ for knowledge by repeated readings, or alternatively, ‘left on the shelf’ by uninterested Readers [31,32]. Readers themselves may experience discomfort hearing directly about others’ distress. The emergent nature of these interactions means that such circumstances cannot be entirely predicted, yet event planners should prepare for and mitigate these eventualities as far as possible by foregrounding implementation strategies (table 4) to promote psychological safety.

### CMO 2 – Humanising others’ experiences

Direct conversations with Books (context) humanise the experience of mental health difficulties (mechanism), contributing to Readers developing a greater empathetic understanding of mental health (outcome).

Event organisers framed direct interaction between Books and Readers as an explicit goal. Many aimed “to bring people together who may not otherwise meet and to also kind of shut down the stereotypes that people have about others” [33]. Events focused on challenging prejudice sought to provide a platform for Books to “show the general public they are ‘human’” [32] and challenge superficial views of important aspects of their experiences and identities by breaking down perceived “us-them” divides between Books and Readers [34]. This could occur through recognition of shared experience including emotional difficulties, to which Readers could relate regardless of personal differences: “Even women [who don’t] look like me, or identify as the same race as me still go through similar struggles. So that was eye-opening because I’ve never really thought about it like that” [35].

This more nuanced understanding of others’ experiences was for some facilitated by the subtle emotional and behavioural cues present in direct interaction, which augmented the interpersonal connection shared by Books and Readers [33,36]. In this context, the experiences and identities Books represent shift from abstract and disembodied concepts to personified and meaningful human experiences [37,38]. Mental health difficulties are thus re-conceptualised from “myths to storied realities” [42], enhancing Readers’ abilities to subsequently understand and empathise with the perspectives of people experiencing similar difficulties [34].

### CMO 3 – Exploring new ways of living

When Living Libraries provide Readers with the permission to explore Books’ experiences through synchronous interaction (context), they will use this opportunity to flexibly explore issues of personal significance (mechanism). This will facilitate awareness of new and helpful ways of living (outcomes).

The synchronous nature of Living Library conversations allows Readers to personalise their interactions by asking questions about aspects of Books’ stories that resonate with their own. As noted by a participant in a mental health-specific event [34], “in a talk, listeners listen passively. As speakers, we are asked to tell our whole story. But in the Human Library, it does not matter whether the sharing is complete or not. The important thing is to let readers know what they want to know.” This form of experiential learning can influence how Readers understand ways of managing their own distress [17] and may help health professionals identify new ways of supporting service users [37,39].

EBCD participants noted that the relative novelty of the Living Library approach and the associated ‘Book’ metaphor can imply that the sharing of experience is intended to be unidirectional, with Readers taking a passive role. Organisers can facilitate interactive dialogue by providing Readers with explicit permission to explore questions of personal significance, within established boundaries. Strategies for reaffirming Readers’ conversational permission include providing library attendees with clear ground rules and example questions or cues to prompt engagement.

### CMO 4 – Personal empowerment

When Living Libraries facilitate the authentic expression of Books’ personal experiences (context), Books will feel that their expertise has been heard and valued (mechanism), contributing to personal empowerment (outcome).

Living Libraries have been used to spotlight marginalised voices [40]. The approach both promotes Books’ sense of having been heard as experts by experience and provides a medium to use their stories to meet personally valued goals, including shaping public attitudes and offering support to others. Personal empowerment therefore emerges from this opportunity for direct self-representation, recognition, and the pursuit of positive change [31,41]. EBCD group members highlighted the potential of a LoLEM to meet the motivation of many experts by experience in mental health to use their unique perspectives to inspire individual, organisational, and social progress against a wider context of stigma and under-recognition. This was mirrored in literature describing participants’ sense of pride and satisfaction after sharing their stories at Living Library events [38,42,43]:

The EBCD group also identified how personal empowerment can occur through meaningful participation in event design and delivery. Extensive involvement of experts by experience was suggested to diminish power imbalances between those with lived experience and organisations, such as health services and universities, that may host a LoLEM. Workshops highlighted that the greater the degree of lived experience involvement and collaborative working alongside staff, the more likely a LoLEM is to reflect the perspective of the groups it seeks to engage. It was suggested that this may reduce the potential for disempowerment emerging from peoples’ stories being misused.

### CMO 5 – Self authoring and co-editing

When Living Libraries support Books with developing a personal narrative to share in conversation with Readers (context), Books will explore and develop insight into these aspects of their lives (mechanism). This leads Books to develop new ways of understanding and sharing their experiences which can contribute to personal growth (outcome).

For Books, authoring and articulating a personal narrative represents a “self-directed process of discovery” [31] and ongoing engagement with story sharing can “demonstrate how their personal identities evolve and develop over time” [43]. This process may contribute to the reframing of mental health difficulties as an aspect of past experience that is of value to the present self [42]. Relatively unstructured dialogue may also facilitate a form of narrative co-editing: “…in the process of creating a narrative in cooperation with readers, books actually alter their understanding of their own self-appointed topic and what it means to them” [31]. Personal growth can therefore result from a shift in the Books’ perceptions of their current circumstances through reflexive story refinement: “Despite occasional moments of discomfort and, perhaps in some cases, because of them, Books recognized that their stories changed because of their participation in the HL [Human Library] Project. Details were added, elements they believed were less important emerged as such, and overall, they achieved greater clarity about their narratives” [35].

Table 3 provides illustrative data supporting each CMO.

**Table 3.** Programme theories with illustrative data extracts.

| Programme theory | Illustrative data | Source |
| --- | --- | --- |
| 1. Psychological safety facilitates dialogue | “[Books’] needs must be recognised and met. This could include things like limiting the number of readings a living book can be required to participate in within a given session, providing adequate support for living books including those with special needs, and providing time and structure for living books to debrief after readings or to read other living books” | [31] |
|  | “ [a living library conversation] will be a deep dive, might need to help these first-time Books prepare e.g. with practice and storytelling and thinking about boundaries” | Workshop 2 Jamboard |
| 2. Humanising others’ experiences | “Readers were given permission to empathize with the experiences of Books and, in the process, see themselves in the Books’ stories” | [35] |
|  | “Stories trigger a response in us that helps us empathise with others” | Workshop 3 field notes |
| 3. Exploring new ways of living | “Another Reader found inspiration in a Book’s journey that he could apply to his own life: “I learned from some people’s experiences that, okay, I’m going through this right now, but you went through it and you overcame it. So I have a choice.” | [41] |
|  | “[Books are] providing other visions/real life examples of ways forward that are hopeful” | Workshop 8 Jamboard |
| 4. Personal empowerment | “Living libraries excel as a strategy for giving voice to marginalised groups. Living library conversations allow for direct self-representation, unmediated by third parties.” | [40] |
|  | “Mental health has been stigmatised for so long, people ignored, locked away and forgotten. Just the very action of telling someone that you want to hear their story is massive.” | Workshop 2 Jamboard |
| 5. Self-authoring and co-editing | “By thinking about and structuring their story, people will often ‘re-author’ their lives, by defining their own existence in relationship to themselves and what they were going through at the time; thereby constructing reality by choices made to give meaning to their lives” | [44] |
|  | “Storytelling can be therapeutic for teller” | Workshop 2 Jamboard |

### Implementation guidance

The integrated realist synthesis and EBCD process contributed to the development of a theory informed implementation guide. Recommendations are summarised here with reference to key stages of event delivery (Table 4). The full guide is available online [28].

**Table 4.**
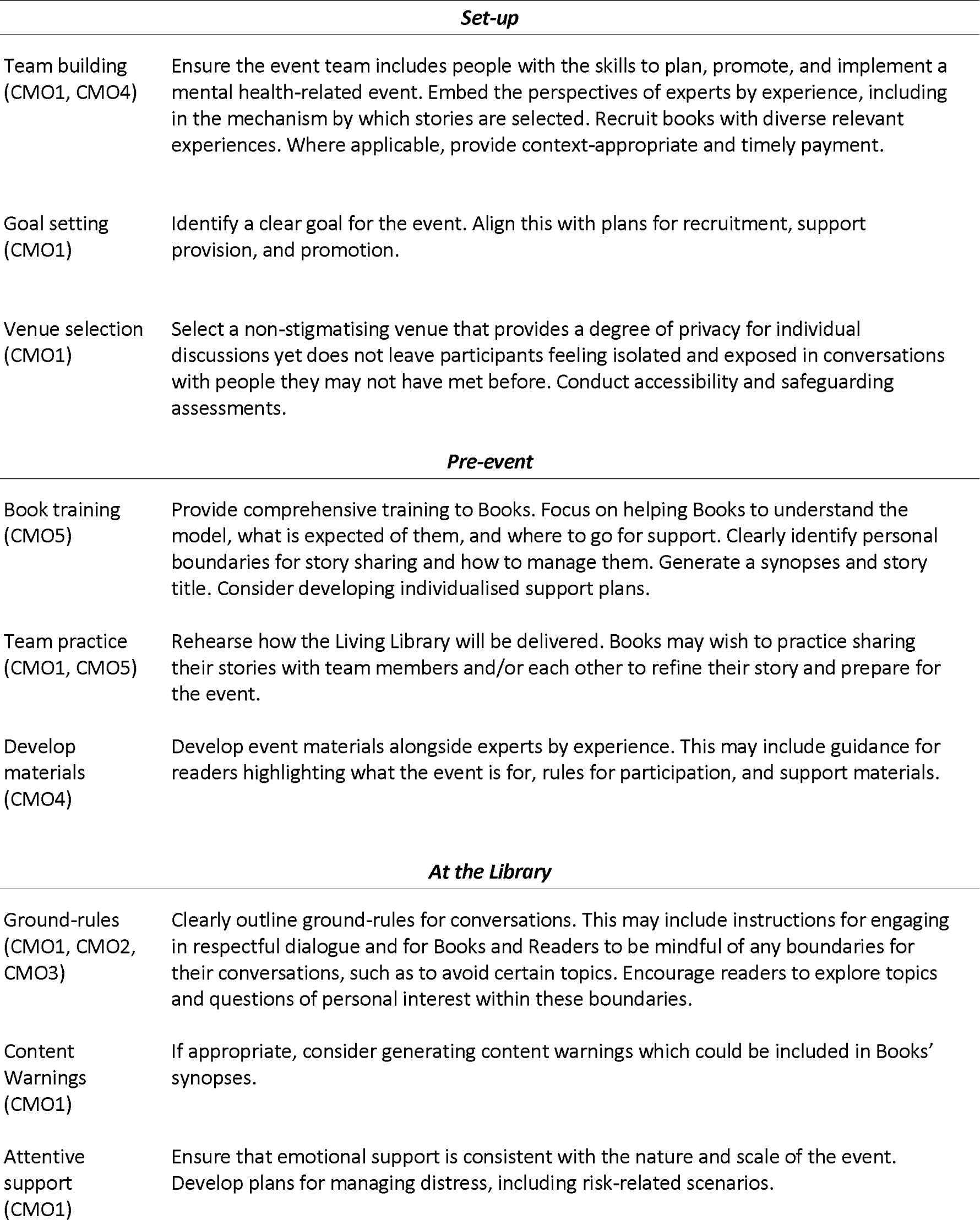
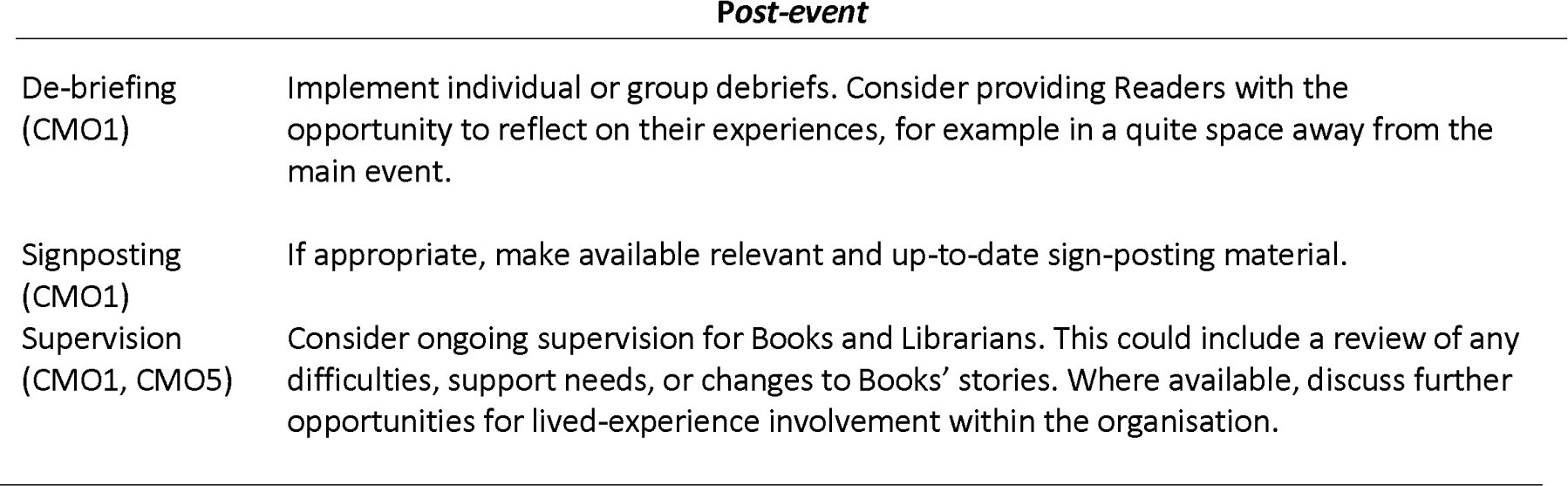
. Co-designed implementation recommendations by event delivery stages.

| *Set-up* |  |
| --- | --- |
| Team building (CMO1, CMO4) | Ensure the event team includes people with the skills to plan, promote, and implement a mental health-related event. Embed the perspectives of experts by experience, including in the mechanism by which stories are selected. Recruit books with diverse relevant experiences. Where applicable, provide context-appropriate and timely payment. |
| Goal setting (CMO1) | Identify a clear goal for the event. Align this with plans for recruitment, support provision, and promotion. |
| Venue selection (CMO1) | Select a non-stigmatising venue that provides a degree of privacy for individual discussions yet does not leave participants feeling isolated and exposed in conversations with people they may not have met before. Conduct accessibility and safeguarding assessments. |
| *Pre-event* |  |
| Book training (CMO5) | Provide comprehensive training to Books. Focus on helping Books to understand the model, what is expected of them, and where to go for support. Clearly identify personal boundaries for story sharing and how to manage them. Generate a synopsis and story title. Consider developing individualised support plans. |
| Team practice (CMO1, CMO5) | Rehearse how the Living Library will be delivered. Books may wish to practice sharing their stories with team members and/or each other to refine their story and prepare for the event. |
| Develop materials (CMO4) | Develop event materials alongside experts by experience. This may include guidance for readers highlighting what the event is for, rules for participation, and support materials. |
| *At the Library* |  |
| Ground-rules (CMO1, CMO2, CMO3) | Clearly outline ground-rules for conversations. This may include instructions for engaging in respectful dialogue and for Books and Readers to be mindful of any boundaries for their conversations, such as to avoid certain topics. Encourage readers to explore topics and questions of personal interest within these boundaries. |
| Content Warnings (CMO1) | If appropriate, consider generating content warnings which could be included in Books' synopses. |
| Attentive support (CMO1) | Ensure that emotional support is consistent with the nature and scale of the event. Develop plans for managing distress, including risk-related scenarios. |
| **Post-event** |  |
| De-briefing<br>(CMO1) | Implement individual or group debriefs. Consider providing Readers with the opportunity to reflect on their experiences, for example in a quiet space away from the main event. |
| Signposting<br>(CMO1) | If appropriate, make available relevant and up-to-date sign-posting material. |
| Supervision<br>(CMO1, CMO5) | Consider ongoing supervision for Books and Librarians. This could include a review of any difficulties, support needs, or changes to Books' stories. Where available, discuss further opportunities for lived-experience involvement within the organisation. |

## DISCUSSION

This integrated realist synthesis and EBCD study generated a programme theory for a LoLEM and theory-informed implementation recommendations. Results emphasise the importance of psychological safety for facilitating productive conversations between Books and Readers. Implementation recommendations highlight ways in which potential organisers may seek to foster supported environments for sharing mental health experiences and further suggestions are reported in a co-designed implementation guide [28].

This study builds on implementation recommendations for the Human Library Organisation’s generalist approach [14]. A notable difference in the context of mental health is the centrality of psychological safety, defined as the ability to take desired interpersonal risks. Occupational research indicates that psychological safety is associated with proactive communication including concern-raising within teams, individual and team-level learning, and work engagement [25]. Recommendations for promoting psychological safety within this literature suggest organisations should seek to create cultures defined by collaboration and interpersonal openness [45]. This aligns with established principles-based approaches to mental health peer-support [46], which point to the significance of safe and trusting relationships within organisations that draw on lived experience, and control over how those experiences are shared. Results here suggest that by investing in training and building meaningful relationships with those sharing their stories, LoLEM organisers can clearly articulate expectations and promote informed personal disclosure. Drawing on established measures [25], further research could evaluate the extent to which these practices influence Books’ perceptions of psychological safety.

Values underpinning mental health peer support, such as mutuality and reciprocity [47], are reflected in evidence of positive impacts amongst those delivering and accessing these services. For example, benefits for those receiving support include improvements in social functioning, hope, and stigma reduction, while peer supporters may experience greater confidence, skills development, and positive reframing of personal identity [48]. Building on this evidence, results of this study suggest that a LoLEM could provide opportunities to learn about ways of understanding and managing mental health from people who may be empowered by being supported to share their stories. Indeed, participants in a Living Library focused on bipolar self-management welcomed the opportunity for direct contact with peers [17], demonstrating the feasibility of using this approach in the peer support context. A LoLEM could also assist service users with sharing stories as part of a recovery-oriented programme of support [34]. Indeed, there is an established body of literature describing the use of storytelling in mental health [6,49] indicating that developing a personal narrative can be highly meaningful for the teller, yet also potentially emotionally challenging [50]. This reinforces this study’s emphasis on dedicated support with story formulation for potential Books.

The LoLEM approach requires adaptation for specific contexts. Organisers should consider factors that could undermine the safe and empowering implementation of lived-experience focused programmes. Survivor research identifies how power imbalance between those with lived experience and health services could serve to discourage perspectives deemed unacceptable to institutional agendas, including critical and marginalised voices, or delegitimize those who draw on their experience in pursuit of systemic change [51]. A recent systematic review of lived experience narratives drew attention to potential misuses of mental health experiences, including commodification, coerced elicitation, harm to narrators or audiences, and curatorial decisions that limit diversity [6]. Good practice recommendations relevant to the LoLEM model include proactive reflection on organisers’ personal biases to ensure power imbalances are not reinforced, encouraging audiences to endorse ethical listening and openness to difference, seeking consent, and promoting control over personal narratives [6]. Consistent with this, findings here emphasise the vital role of lived experience perspectives in implementing transparent and collaborative LoLEMs. Established models for involvement, including EBCD, could be used to facilitate development of future programmes.

### Implications for research

Further work may explore the feasibility of Living Libraries as sustained programmes and whether effects persist over time. CMOs reported here could inform the selection of outcomes. The humanising impact of a LoLEM would be expected to contribute to more empathetic perspectives on mental health and in the context of peer-support, future research may investigate potential impacts on Reader self-efficacy. For Books, the proposed positive impact of storytelling and personal empowerment aligns with definitions of personal recovery for which several assessments have been developed [52]. Factors underpinning differences in outcomes across varied service settings, and how this model may fit alongside existing lived experienced-based interventions, are currently unknown. These issues could be addressed using realist evaluation methodology.

### Implications for practice

While lived experience opportunities continue to grow within health services, most roles do not involve training in self-disclosure [53]. The LoLEM model could bridge the gap between people with mental health experiences seeking opportunities to help others, and more formal paid positions requiring considerable time commitments. The approach could also complement efforts to increase lived experience expertise in health professional training, as evidenced by its successful implementation in social work training [37]. More specifically, a LoLEM could afford healthcare trainees opportunities to engage with a broader range of people than they may typically meet, and in neutral settings outside of clinical environments which may be more conducive to open conversation. It is also feasible to combine a Living Library with structured reflexive activities to embed learning and consider the applicability of a Book’s story to a Reader’s clinical practice [39]. Actions to promote equality, inclusion and diversity in the UK health service should apply an intersectional lens [54]. It has been noted that the Living Library model is well suited to assist health professional trainees with developing a lived appreciation of intersectionality through conversations that illuminate how aspects of identity interact [35]. As per the programme theory reported here a LoLEM could serve to humanise and promote the understanding of nuanced relationships between mental health experiences and other salient aspects of identity.

### Strengths and limitations

A strength of this approach is the breadth of mental health experience represented in the study team. PPI perspectives were sought throughout, including during study design, via an advisory group and in co-design meetings, and to assist with dissemination. This facilitated the generation of findings that reflect a variety of perspectives and contexts in which a LoLEM could be hosted. This project also demonstrates the feasibility of a novel and creative approach to mental health programme co-design and theory generation. However, this study does have limitations. Many evidence sources identified in systematic searches were non-mental health specific and involved short-term evaluations. This study principally recruited from the North of England which may limit the transferability of its findings. In a change to the study protocol [21], theory refinement interviews [23] were not conducted due to limitations in available resources. The reported search on psychological safety was similarly limited to a single multidisciplinary database due to a limited capacity to manage the screening burden of a larger search.

## CONCLUSION

This project is the first to apply realist methods to articulate explanations for how Living Libraries may work and adds to existing literature by identifying specific strategies for promoting the safety and effectiveness of this model as applied to mental health experiences. Continued recognition of the importance of lived experience in shaping mental health policy and practice justifies further consideration of how this approach can be implemented and evaluated.

## PUBLIC AND PATIENT INVOLVEMENT

Experts by experience in mental health contributed to all stages of this study. A stakeholder group informed the initial study design and experts by experience were co-applicants for funding. The study team included a service user research and experts by experience were involved in an advisory group, all of whom are involved in authoring key study outputs. The co-design group involved service users and carers and was led by facilitators who delivered workshops from a lived experience perspective. Participants and team members with lived experience assisted with dissemination at local and national events.

## ETHICS APPROVAL

This study involved human participants and granted ethical approval by Coventry and Warwick National Health Service Research Ethics Committee on 29th December 2021 (REF: 305975). Participants gave informed consent to participate in the study before taking part.

## DATA AVAILABILITY STATEMENT

No data are available.

## AUTHOR CONTRIBUTIONS

FL designed and led the study. PM conducted data collection and analysis for the realist synthesis. JB supported search strategy development. KM and GC co-facilitated EBCD workshops with support from SJ, RJ, ZG, CL, PJ and LW. JR-M provided methodological guidance for the realist synthesis. SF, CI, HK, SP, EM, and MS formed an expert advisory group which provided strategic guidance across the study. SR supported article writing. All authors provided approved the final article.

## TWITTER

Steven Jones @lancsspectrum

Sarah Powell @DrSarahPowell1

John Barbrook @stackingbooks

Paul Jebb @pauljebb1

Lesley Whittaker @Lesleyp222

## ACKNOWLEDGMENTS

The research team would like to thank all those who contributed to the co-design of the outputs for this study.

## APPENDIXES

## Appendix A Initial programme theories

### 1. Readers – hearing stories about mental health

#### Intended impacts

1.1 Reduced isolation - If readers hear stories that resonate with their own experiences of mental health difficulties, then they will feel that their experiences are shared, thus normalising their experience and reducing their sense of social isolation. This shared experience may make them feel they are more likely to be understood by the book, and by others with similar experiences, and therefore more able to talk about their mental health difficulties with others, promoting social engagement and help-seeking.

1.2 Tailored learning - If readers take part in a library that facilitates synchronous interaction, then they will be able to ask the questions about mental health that they want answered, increasing the relevance of the information shared by the book to their personal experience and individual learning needs. This will mean readers are better informed and have the information they need to manage their own mental health difficulties.

1.3 Challenging preconceptions - If readers interact with a living book who through their participation in the library challenges the reader’s negative preconceptions or stigma regarding mental health, for example that it is not possible to live well with a mental health problem, then contact with the book will disconfirm these preconceptions, increasing hope for their own recovery.

1.4 Connecting to the individual - If the reader engages with a book in a synchronous, face-to-face interaction, then this intimate interpersonal context facilitated will promote empathy with the book as their storytelling will be perceived as more authentic and emotive, facilitating other intended impacts (such as learning, social connection, and hope for recovery).

1.5 Connecting to the story - If the reader interacts with a book whose story involves a compelling narrative about their mental health difficulties, then the reader will be more engaged by their story, making the story more likely to be internalised and remembered, facilitating other impacts such as lasting change in perspective. This impact will be greater in a context where shared life experiences create personal resonance with the story.

#### Unintended impacts

1.6 Exposure to distressing stories - If the reader interacts with a book who recalls distressing aspects of their experiences and/or who becomes distressed when doing so, then the reader may be at increased risk of themselves becoming distressed.

1.7 Interpersonal tension - If the reader engages with a book who they perceive holds views that they find objectionable, then there is an increased risk of interpersonal tension and possible subsequent distress is increased The reader is also unlikely to identify with the book, and more likely to actively reject any strategies or ideas that the book offers, so will not learn anything of value and feel they have wasted their time. This could make them less likely to engage with other forms of peer-based support in future.

1.8 Negative social comparison - If the reader engages in negative social comparison, for example perceives that they are unlikely to achieve the level of recovery experienced by the living books at the library, then they may feel dejected and less hopeful about their own recovery, and therefore less likely to engage in recovery focussed activities. This is particularly likely to happen if the books are selected for “success” (or “recovery”) stories only.

### 2. Living ‘books’ – telling stories about mental health

#### Intended impacts

2.1 Using experiences to help others - If living books tell their stories of mental health difficulties to readers who they feel have been derived benefit from hearing their experiences, those telling their stories will derive satisfaction from their participation contributing to a sense of personal meaning.

2.2 Developing personal understanding - If living books tell their stories of mental health difficulties, then the process of recalling and communicating their story in a safe environment may facilitate the development of new understandings of their previous experiences, contributing to greater awareness of their own mental health.

2.3 Social inclusion - Where living books are encouraged to use their mental health experiences as part of a mental health programme to help others, books may begin to value and drawn on these experiences to achieve personal goals such as greater social engagement, thus promoting social inclusion and involvement in other activities that invite them to draw on their lived expertise.

2.4 Being empowered - If living books are supported to tell their stories of mental health difficulties in a way that is authentic to their experience and aligns with their motivation for sharing their story, then they will experience a sense of empowerment because the process of storytelling will promote autonomy over expression of an experience that in society more broadly is often stigmatised and supressed.

#### Unintended impacts

2.5 Recalling distressing experiences - If living books tell their experience of mental health difficulties, without adequate training and support, then the process of recalling painful experiences may trigger distress because they are still emotionally processing these experiences.

2.6 Devaluing of lived experiences - If living books attempt to tell their experiences to a reader who is perceived to be disinterested, disrespectful or insensitive, then they will be at increased risk of feeling that their lived experiences of distress have devalued contributing to disengagement from the library.

### 3. Organisation and support

3.1 Balancing conversational openness and risk of harm - Impacts of a living library for mental health will be moderated by organisational aims, including the purpose of the library, and organisational culture, including the extent to which lived experience of distress is deemed to be an important resource for shared learning. These factors will determine how organisers, or ‘librarians’, understand the need to balance the facilitation of open conversations about mental health with the need to restrict or structure conversations to reduce perceived risk to participants. This will determine when and how they intervene to support conversations within the library.

3.2 Curation and commodification - If the stories selected for the library, and the way they are told (curation), is perceived by books to be prescriptive to the extent that their stories are no longer authentic expressions of their lived experience, then books may perceive that their stories of mental health experiences are being commodified – used for a purpose that is incongruent with their motivation for participating in the library - and thus devalued. This is likely to lead to them withdrawing participation in the library.

3.3 Location facilitates engagement - If the physical location in which the library is held facilitates a feeling of psychological safety, for example within a space that means personal disclosure remains private and where participants are not overlooked by the public, then both books and readers will be more likely to engage in open and engaging conversations because they will feel better able to immerse themselves in conversation without distraction or concern over confidentiality.

3.4 Perceived emotional support - Where books and readers perceive that the library provides or will provide sufficient emotional support to address any potential distress, they will feel safe and held, and so and will be more open to engaging with potentially distressing conversations about mental health.

3.5 Setting expectations - Impacts of a living library are likely to depend on the expectations of both books and readers. Where it is made clear to participants what is expected of them within the library and any boundaries in place, participants will be better equipped to manage any difficulties that may arise and/or engage with the programme as intended, because they will understand the purpose and function of the library.

3.6 Setting ground rules – Where the ‘librarians’ or organisers set clear guidelines for engaging in conversations about mental health, including the necessity of communicating respectfully and what to do if difficult conversation do arise, both books and readers will feel empowered to engage in more open and sincere conversation.

## Appendix B Targeted search for theories related to psychological safety

This search was conducted in June 2022 on Web of Science. Resources available for the review precluded full review of the research literature and as such, a large multi-disciplinary database was selected for this search. Its aim was to identify English language articles in health-related research that included middle range theories related to the concept of psychological safety. Search terms were informed by the BeHEMoTh framework [24] and combined with the ‘AND’ Boolean operator:

**Be**haviour of interest: “psychological safety”

**H**ealth context: “health”

**E**xclusions: n/a

**M**odels or **th**eories: “model* or theor* or concept* or framework*”

The search returned 290 articles. Title and abstracts were screened by PM. Articles that appeared relevant to the research question and suggested that they may include a middle range theory were flagged for full-text screening. Twenty-nine articles were read by PM and RJ who held collaborative discussions, through which one article was identified as including a middle range theory of psychological safety relevant to the developing programme theory [25].

## Appendix C Characteristics of included studies

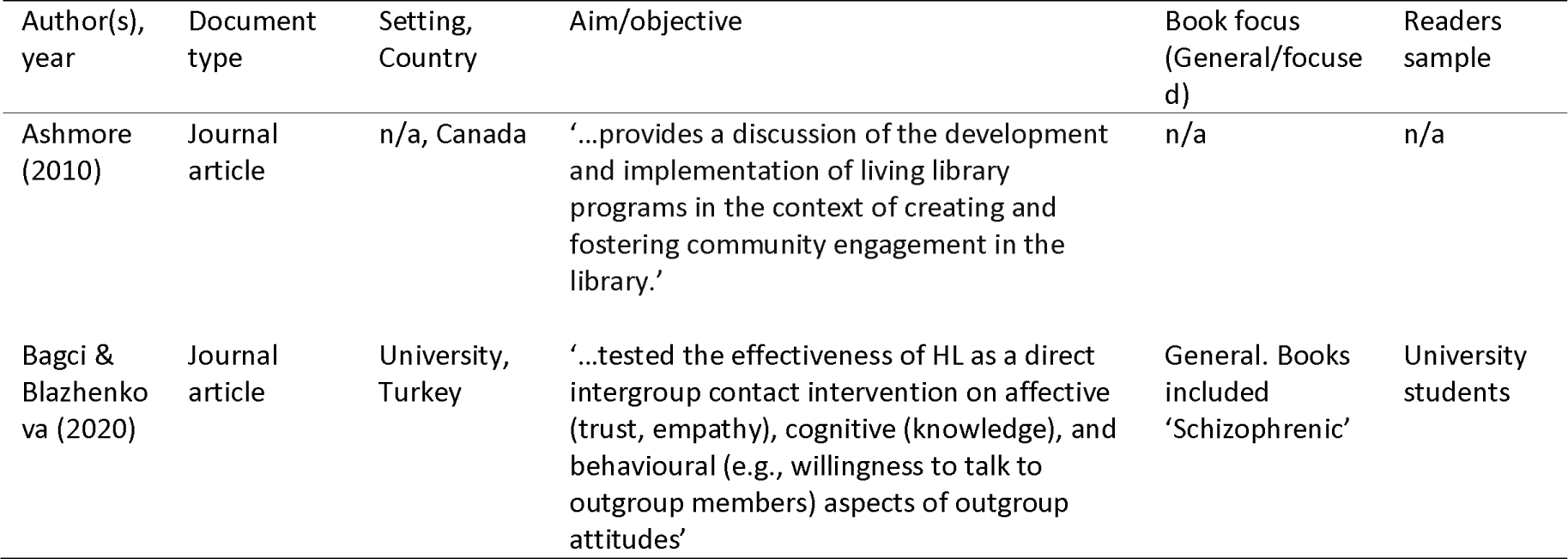
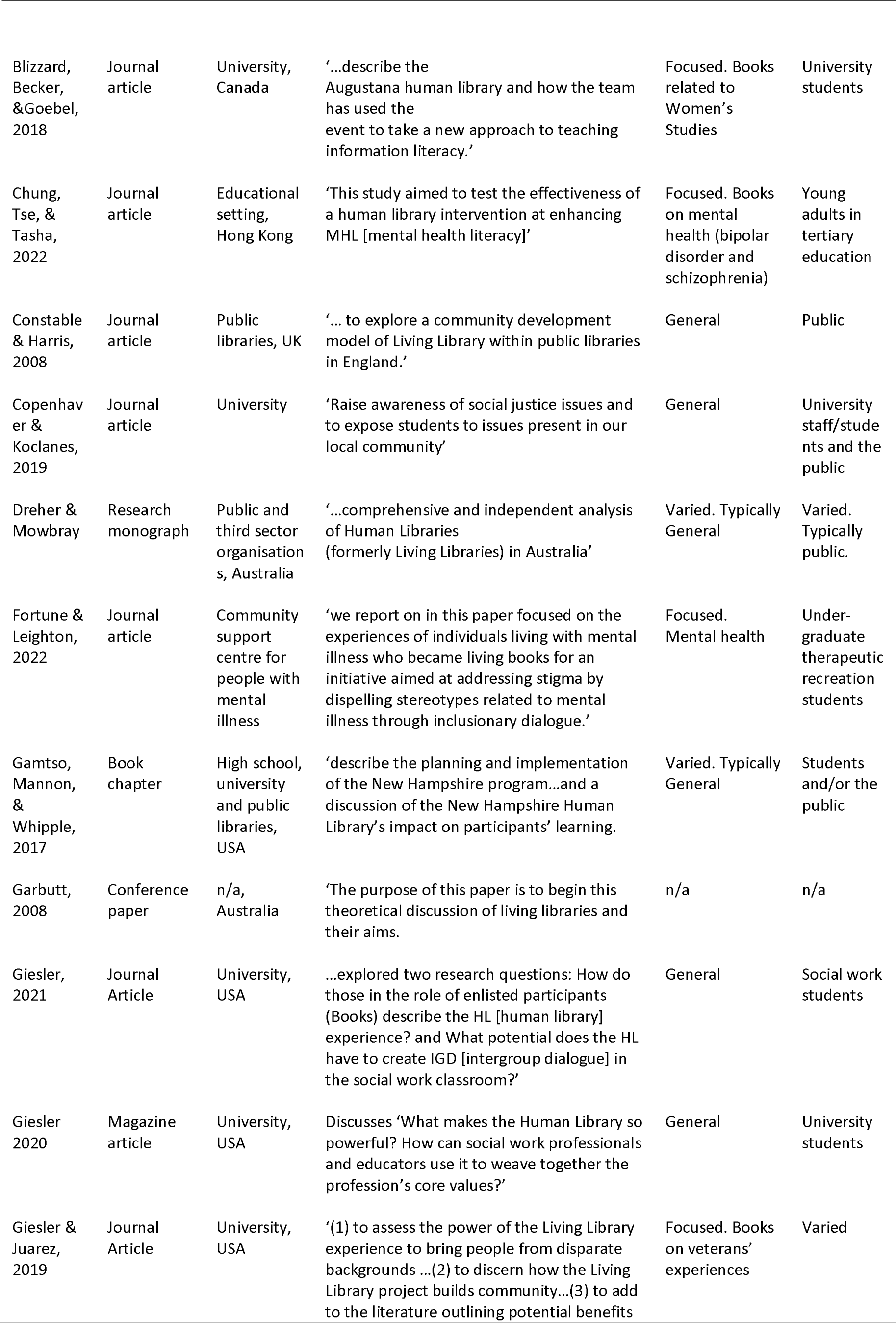
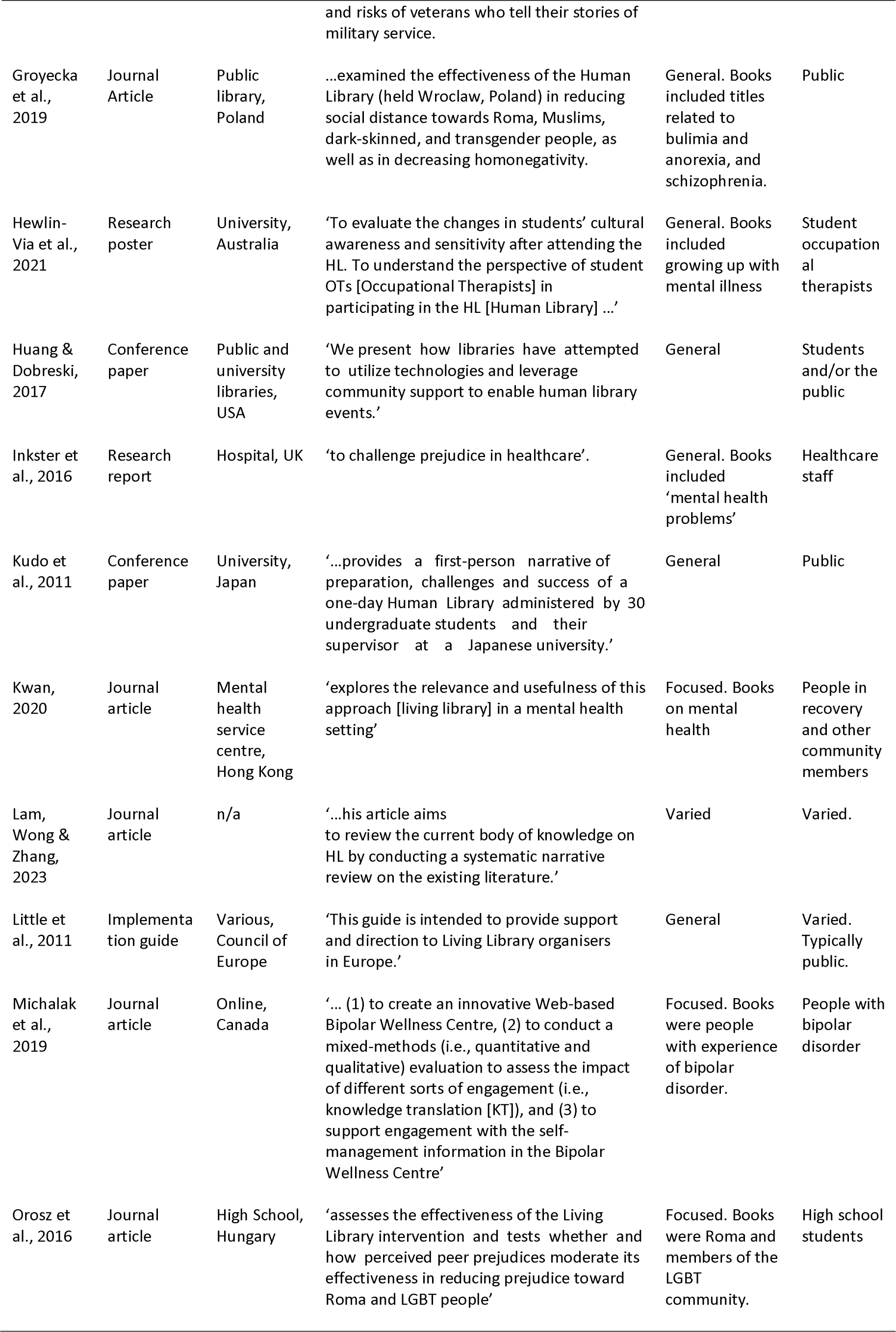
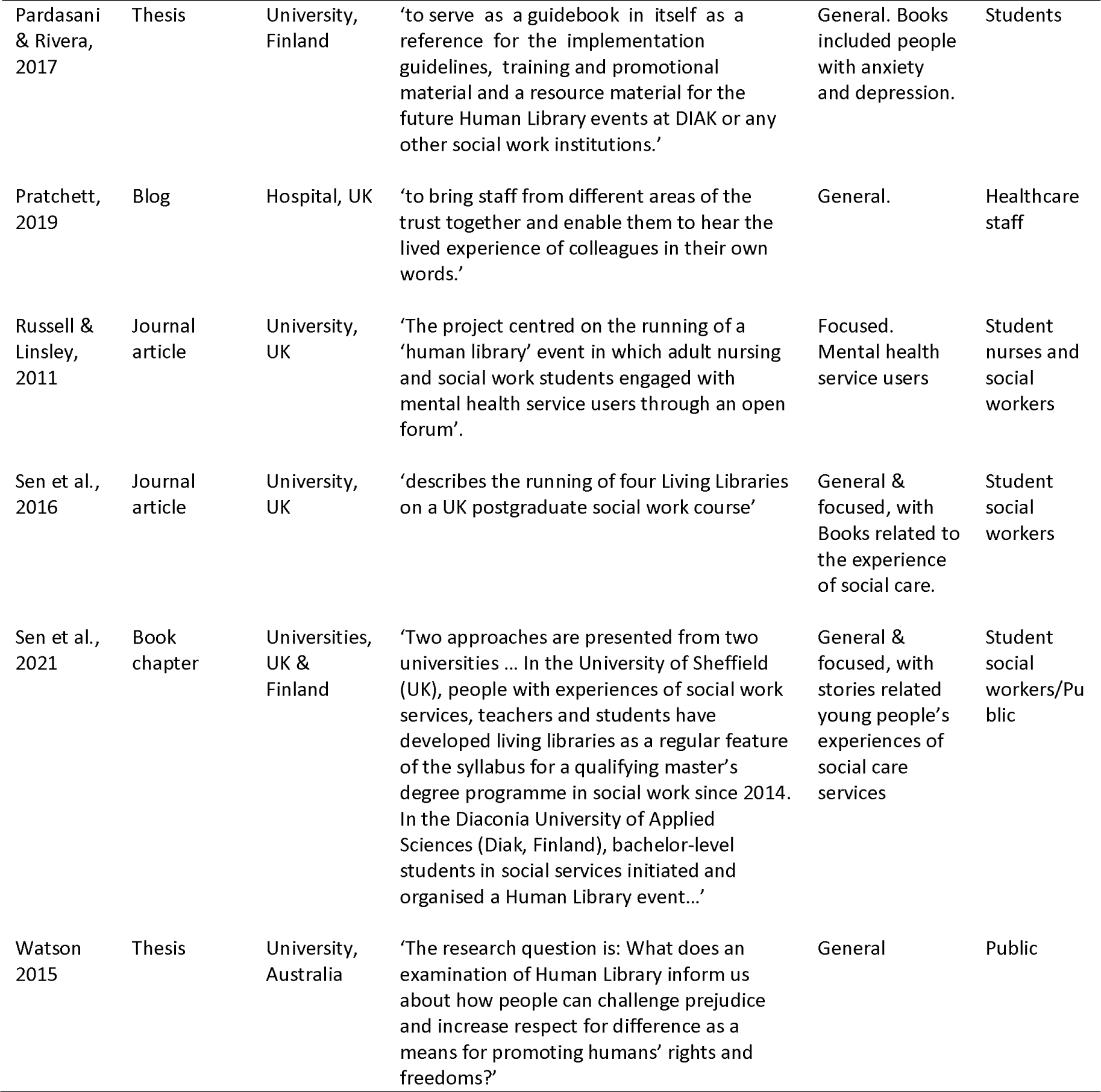

